# The Prevalence of Sexual Harassment in Internal Medicine in Switzerland

**DOI:** 10.1101/2025.08.27.25334541

**Authors:** Olivia Wassner, Christa Nater, Jeanne M. Barbier, Sven Streit, Jeanne Moor

**Author notes:** Co-senior authors (Sven Streit, Jeanne Moor).

## Abstract

**Background:** Sexual harassment (SH) impairs physician well-being and performance, yet data on the epidemiology of SH in Internal Medicine are scarce. Here, we set out to assess the frequency and patterns of SH experienced by physicians in Switzerland.

**Methods:** We performed secondary analyses of a web-based survey that focused on physicians’ and medical students’ well-being and career ambitions but also asked for past experiences of verbal and physical SH. Specifically, we asked if they had ever experienced SH at the workplace, and if yes, how often they experience SH. We then conducted gender-stratified descriptive analyses of sexual harassment frequency and determined type and gender of the perpetrators from both close- and open-ended questions. We used Chi-square or Fisher’s Exact test for comparisons across genders.

**Results:** Respondents consisted of 682 physicians (59% women, mean age of men 36, standard deviation [SD] of 10 years, mean age of women 39, SD of 12) and 321 medicals students (70% women, mean age of 25, SD of 4 in both gender groups). Among the physicians, verbal SH was reported by more female than male physicians (74% vs. 38%, p<0.01 for gender difference) and by more female than male medical students (56% vs. 38%, p=0.04). Physical SH was also reported by more female than male physicians (34% vs. 15%, p<0.01) and by more female than male medical students (24% vs. 9%, p=0.01. For both women and men victims, perpetrators were predominantly physicians and patients of the opposite sex.

**Conclusion:** This study provides evidence for a high prevalence of both verbal and physical SH experienced by both physicians in Internal Medicine and medical students in Switzerland, particularly among the women. Existing barriers to reporting and prevention of SH should be identified to improve healthcare work environments in Switzerland.

## Introduction

Sexual harassment (SH) in medicine affects the wellbeing and work performance of physicians and has become a focus of media attention in Switzerland (1–3). In 2022, a survey targeting female physicians from all specialties in Switzerland had 252 respondents indicating that they experienced SH at their workplace (4). Internal medicine seems to be particularly affected. A U.S. study found that one in four female internal medicine residents has experienced unwanted sexual attention (5).

Data specifically collected on SH in internal medicine have not previously been reported in Switzerland. A Swiss study from 2015-16 including physicians from all specialties found a comparably low prevalence of 0.7% (6). Yet, a more recent study conducted in 2020, including junior doctors from all disciplines in the canton of Vaud, found that 44% of respondents reported having experienced or witnessed sexist remarks or verbal SH (7). For medical students, the prevalence of SH experiences was found to be 16% in a study conducted in 2021 among 1033 medical students in the French-speaking part of Switzerland (8). As most victims of SH are women, prevalence of SH is higher for women compared to men (9– 11).

Comprehensive data on the impact of SH on physicians is rare. However, other professions such as nurses show that SH is related to psychological and physical health issues as well as emotional distress (12). Furthermore, SH decreases workplace satisfaction and can worsen work performance (13).

To address the current lack of information on prevalence in internal medicine in Switzerland, we conducted a cross-sectional survey among Swiss internal medicine physicians and medical students. Additionally, we asked about perpetrator type. In line with previous literature on SH, we analyzed a previously conducted cross-sectional quantitative web-based survey.

## Methods

The analyzed survey focused on examining the wellbeing and career aspirations of physicians in general internal medicine. The survey consisted of questions about career ambitions, physician well-being, and workplace related factors such as gender-related discrimination but also two questions about sexual harassment. Results from this survey on the original research questions have been addressed elsewhere (14).

### Study design, setting and participants

The cross-sectional web-based survey was provided to Swiss general internal medicine physicians and medical students in the 4^th^ to 6^th^ year of their studies at the University Bern. Clinical internships start in the 4^th^ year of studies, so medical students from the preclinical part of education (years 1 through 3) were not included. To reach physicians, the survey was sent to the heads of departments or administration of 14 Swiss hospitals and 6 ambulatory or primary care institutes who distributed it to their employees via email. A link to the survey was posted in the newsletter of the Swiss society of general medicine. Further, the Swiss young general practitioners’ association sent an email with the link to the survey to their members. We also placed an advertisement in the Swiss journal *Primary Hospital Care*.

The survey was available online between December 2021 to April 2022. Participants consented to anonymous participation. The ethics committee of the canton of Bern waived this study (identifier Req-2021-01085).

### Data collection

The survey was administered on the platform www.surveymonkey.com. We collected the following general demographic information: age, self-reported gender, language (German, French or Italian), workplace type (university hospital, other hospital, private practice with <3 or >3 colleagues), and current job position.

To self-identify their gender, participants were given the options of woman, man and diverse.

### Outcomes

We sought to establish the prevalence of verbal and physical SH, providing a definition embedded in the question. The survey asked participants to rate how often they experienced verbal or physical SH on a Likert scale: never; less than once a year; once or twice a year; three to five times a year; once or twice a month; weekly; daily.

We asked the following question to establish the prevalence of verbal harassment:

a. Have you experienced sexual harassment in the form of lewd remarks (e.g. about your own appearance) during your internships at the hospital or clinical work? To establish the prevalence of physical SH we asked the following question :
b. Have you experienced sexual harassment in the form of unwanted physical contact during your internships at the hospital or clinical work?

Additionally, the participants that reported sexual harassment were given the option of describing the perpetrators of verbal or physical SH. They could choose one or multiple perpetrator types out of the following: female physician; male physician; female patient; male patient; other. For the option of *other* participants were prompted to describe the perpetrator in free text.

The free text answers were structured into the following categories by two independent reviewers (OW, JB): nurse (no gender specified); female nurse; male nurse; other; unknown. Responses that fell into the category *unknown* were excluded, as these answers did not describe a person.

### Survey development and testing

We piloted the full questionnaire on a convenience sample of ten physicians. We collected feedback about the duration of the survey, coherence and completeness of questions and technical functioning of the online survey. The reviewers reported that it took 12 minutes to fill out the survey, and despite its length it was perceived as well constructed and addressing important topics. Native speakers translated the final questionnaire back and forth, from English to German or French and then back to English to ensure coherence (15). Participants could choose to fill out the online survey in German or French. We include those and the English version in our web supplement.

### Statistical analysis

Descriptive statistical analysis included participation percentages for physicians and students, as well as prevalence of SH and the frequency of harassment experiences. We performed Chi square tests or Fisher’s Exact test in R statistics (version R 4.5.0). The threshold of significance was set at two-sided p < 0.05.

## Results

### Basic demographic of respondents

A total of 682 physicians and 321 medical students completed the survey. The sample of physician respondents represents 8% of the total population of physicians that worked in general internal medicine in Switzerland in 2022 (N = 8511 (16)). The sample of students represents 39,6% of medical master students at the University of Bern in the fall semester of 2021 (N = 811) (17). Of physician respondents, 59.2% were female and 70.4% of student respondents were female. Two physicians self-identified their gender as diverse. To assure their anonymity, we excluded their responses from this study, as reporting their demographic data might reveal their identity. The mean age of male physicians was 39 (SD 12) years and 36 (10) years in female physicians (Table 1). The biggest part of physicians was from the German speaking part of Switzerland and worked in university or other hospitals (Table 1). Among medical students, mean age was 25 (SD 4) years in both women and men (Table 2). Most of the students were in their fifth year of study. Of medical student respondents, 29,7% were men and 70,3% were women. None of the student respondents were diverse.

**Table 1:** Physicians’ Characteristics.

| Characteristic | Female physicians<br>N=404 |  | Male physicians<br>N=278 |  |
| --- | --- | --- | --- | --- |
| Age in years, mean (SD) | 36 (10) |  | 39 (12) |  |
| Swiss language region |  |  |  |  |
| German-speaking | 288 | 71% | 197 | 71% |
| French-speaking | 111 | 27% | 73 | 26% |
| Italian-speaking | 5 | 1.2% | 8 | 2.9% |
| Type of workplace |  |  |  |  |
| University hospital | 155 | 38% | 104 | 37% |
| Other hospital | 152 | 38% | 115 | 41% |
| Private practice with <3 colleagues | 36 | 8.9% | 29 | 10% |
| Private practice with ≥3 colleagues | 43 | 11% | 23 | 8.3% |

**Table 2:** Medical Students’ Characteristics.

| Characteristic | Female students<br>N=226 | Male students<br>N=95 |
| --- | --- | --- |
| Age in years, mean (SD) | 25 (4) | 25 (4) |

| Study year |  |  |  |  |
| --- | --- | --- | --- | --- |
| 4th | 51 | 23% | 20 | 21% |
| 5th | 94 | 42% | 42 | 44% |
| 6th | 81 | 36% | 33 | 35% |

### Prevalence of verbal SH

#### Female physicians and medical students

Of female physicians, 74% (n=272) reported verbal SH; 9.8% (n=36) indicated they had been verbally sexually harassed more than once per month (Table 3). Of female students, 56.2% (n=118) reported verbal SH; 11.5% (n=24) indicated that they were verbally sexually harassed more than once a month (Table 3).

**Table 3:** Prevalence and gender differences of verbal sexual harassment.

|  | female physician<br>(N = 404) |  | male physician<br>(N = 278) |  | female student<br>(N = 226) |  | male student<br>(N = 95) |  |
| --- | --- | --- | --- | --- | --- | --- | --- | --- |
| never | 94 | 25.7% | 152 | 62.3% | 92 | 43.8% | 56 | 63.6% |
| <1 per year | 109 | 29.8% | 42 | 17.2% | 35 | 16.7% | 12 | 13.6% |
| 1-2 per year | 79 | 21.6% | 28 | 11.5% | 40 | 19.0% | 14 | 15.9% |
| 3-5 per year | 48 | 13.1% | 15 | 6.1% | 19 | 9.0% | 2 | 2.3% |
| 1-2 monthly | 24 | 6.6% | 7 | 2.9% | 22 | 10.5% | 3 | 3.4% |
| weekly | 12 | 3.3% | 0 | - | 2 | 1.0% | 1 | 1.1% |
| unknown | 38 |  | 34 |  | 16 |  | 7 |  |
| Chi-square test: Physicians $p < 0.001$. Students $p = 0.04$ | | | | | | | | |

**Table 4:** Prevalence and gender differences of physical sexual harassment.

|  | female physician<br>(N = 404) |  | male physician<br>(N = 278) |  | female student<br>(N = 226) |  | male student<br>(N = 95) |  |
| --- | --- | --- | --- | --- | --- | --- | --- | --- |
| never | 241 | 66.2% | 207 | 84.8% | 160 | 76.2% | 80 | 90.9% |
| < 1 x per year | 95 | 26.1% | 32 | 13.1% | 29 | 13.8% | 4 | 4.5% |
| 1-2 per year | 20 | 5.5% | 5 | 2.0% | 20 | 9.5% | 3 | 3.4% |
| 3-5 per year | 5 | 1.4% | 0 | - | 0 | - | 1 | 1.1% |
| 1-2 per month | 2 | 0.5% | 0 | - | 1 | 0.5% | 0 | - |
| weekly | 1 | 0.3% | 0 | - | 0 | - | 0 | - |
| unknown | 40 |  | 34 |  | 16 |  | 7 |  |

Chi-square test: Physicians $p < 0.001$ . Students $p = 0.01$ (never vs. $< 1x/year$ vs. $\geq 1x/year$ )

**Table 5:**
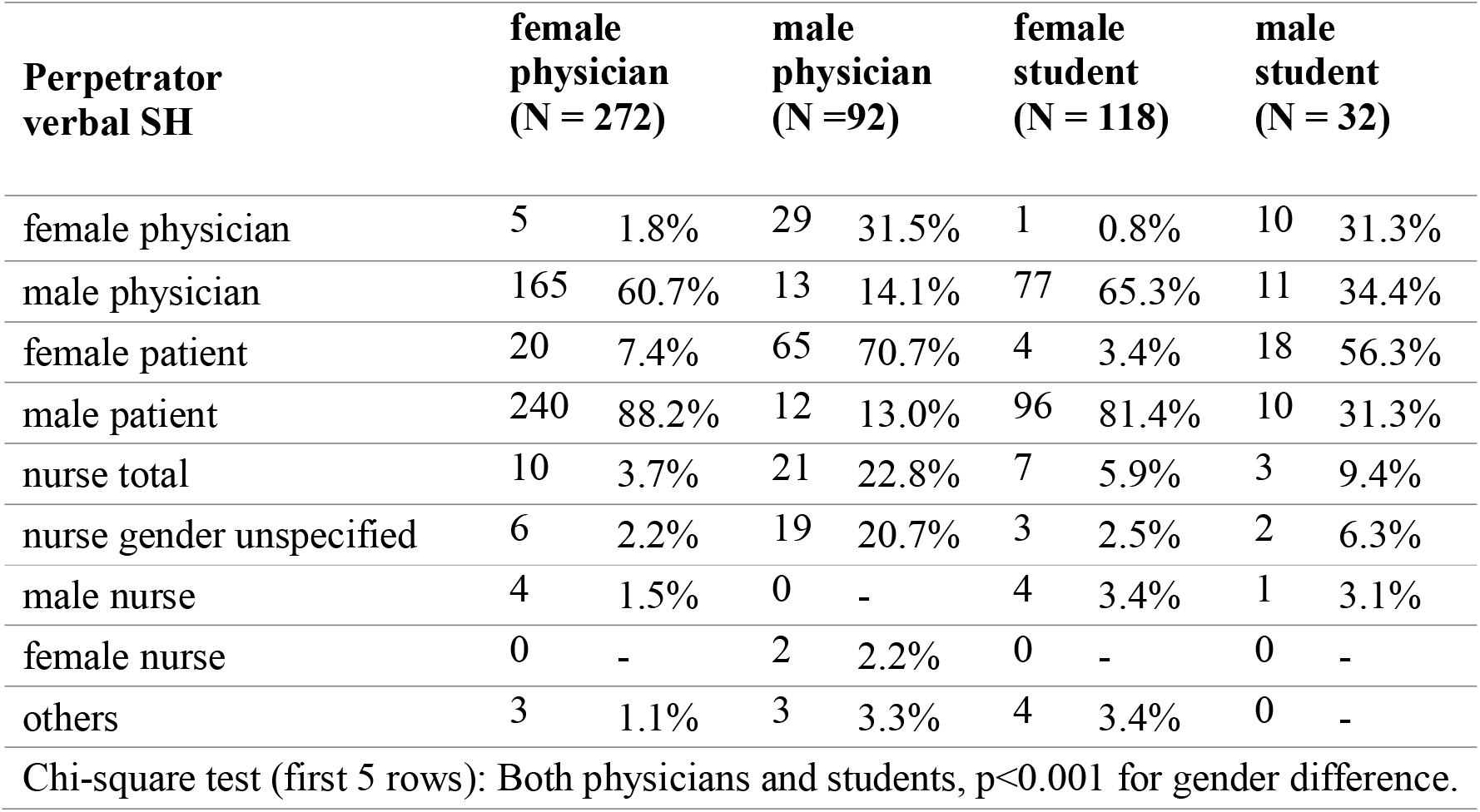
Reported perpetrator by respondents for sexual harassment.

| **Perpetrator verbal SH** | **female physician (N = 272)** |  | **male physician (N = 92)** |  | **female student (N = 118)** |  | **male student (N = 32)** |  |
| --- | --- | --- | --- | --- | --- | --- | --- | --- |
| female physician | 5 | 1.8% | 29 | 31.5% | 1 | 0.8% | 10 | 31.3% |
| male physician | 165 | 60.7% | 13 | 14.1% | 77 | 65.3% | 11 | 34.4% |
| female patient | 20 | 7.4% | 65 | 70.7% | 4 | 3.4% | 18 | 56.3% |
| male patient | 240 | 88.2% | 12 | 13.0% | 96 | 81.4% | 10 | 31.3% |
| nurse total | 10 | 3.7% | 21 | 22.8% | 7 | 5.9% | 3 | 9.4% |
| nurse gender unspecified | 6 | 2.2% | 19 | 20.7% | 3 | 2.5% | 2 | 6.3% |
| male nurse | 4 | 1.5% | 0 | - | 4 | 3.4% | 1 | 3.1% |
| female nurse | 0 | - | 2 | 2.2% | 0 | - | 0 | - |
| others | 3 | 1.1% | 3 | 3.3% | 4 | 3.4% | 0 | - |

Chi-square test (first 5 rows): Both physicians and students, $p < 0.001$ for gender difference.

**Table 6:** Reported perpetrator by respondents for sexual harassment.

| **Perpetrator of physical SH** | **female physician (N = 123)** |  | **male physician (N = 37)** |  | **female student (N = 50)** |  | **male student (N = 8)** |  |
| --- | --- | --- | --- | --- | --- | --- | --- | --- |
| female physician | 1 | 0.8% | 9 | 24.3% | 0 | - | 2 | 25.0% |
| male physician | 59 | 48.0% | 4 | 10.8% | 28 | 56.0% | 4 | 50.0% |
| female patient | 2 | 1.6% | 21 | 56.8% | 1 | 2.0% | 6 | 75.0% |
| male patient | 72 | 58.5% | 3 | 8.1% | 26 | 52.0% | 2 | 25.0% |
| nurse total | 2 | 1.6% | 5 | 13.5% | 0 | - | 0 | - |
| nurse gender unspecified | 1 | 0.8% | 4 | 10.8% | 0 | - | 0 | - |
| male nurse | 1 | 0.8% | 0 | - | 0 | - | 0 | - |
| female nurse | 0 | - | 1 | 2.7% | 0 | - | 0 | - |
| others | 3 | 2.4% | 2 | 5.4% | 2 | 4.0% | 0 | - |

Chi-square test (first 5 rows): Physicians $p < 0.001$ for gender difference.

#### Male physicians and medical students

Of male physicians, 37.7% (n = 92) reported they had been verbally sexually harassed at least once (Table 3). Of male students, 36.4% (n=32) reported they had been verbally sexually harassed at least once; 4.5% (n=4) noted that they were verbally sexually harassed more than once a month.

### Prevalence of physical SH

#### Female physicians and medical students

Physical SH was less commonly reported than verbal SH. Of female physicians, 33.8% (n= 123) reported having been physically sexually harassed; 0.8% (n=3) indicated they had been physically sexually harassed more than once a month. Of female students, 23.8% (n=50) reported having been physically sexually harassed. One female student (0.5%) indicated had been physically sexually harassed once or twice a month.

#### Male physicians and medical students

Male physicians reported physical SH half as frequently (15.2%; n=37). No male physicians reported being physically sexually harassed more than once a month.

Of male students, 9.1% (n=8) reported they had been physically sexually harassed. No male student reported being physically sexually harassed more than three to five times per year.

Prevalence of verbal and physical SH significantly differed as a function of gender (verbal: p =0.04; physical: p =0.01).

### Perpetrators of verbal and physical SH

#### Female physicians and medical students

For female physicians, male patients were the most common perpetrators of verbal harassment (88.2%) and physical harassment (58.5%). Of female physicians, 60.7% indicated that they were verbally sexually harassed by a male physician; 48% reported they had been physically sexually harassed by a male colleague. Female physicians were verbally sexually harassed by nurses in 3.7%; 1.6% of female physicians were physically sexually harassed by nurses.

Female students reported that male patients (81.4%) and male physicians (65.3%) were the most frequent perpetrators of verbal SH; 56% of female students reported they had been verbally sexually harassed by male physicians; 52% reported they had been verbally sexually harassed by male patients.

#### Male physicians and medical students

Of male physicians who reported they had been harassed, 70.7% reported being verbally sexually harassed by female patients; 31.5% reported they had been physically sexually harassed by female physicians. In 56.8% of cases male physicians reported having been physically sexually harassed by female patients; 24.3% reported they had been physically sexually harassed by female physicians. Additionally, 22.8% of male physicians reported having been verbally sexually harassed by nurses (female or male), while 13.5% reported they had been physically sexually harassed by a nurse.

Of male students, 56.3% reported they were verbally sexually harassed by female patients, while 34.4% of male students reported having experienced verbal SH by male physicians. Male students reported in 75% of physical SH cases that female patients were perpetrators, and 50% of male students cited male physicians as perpetrators of physical SH. In summary, the majority of perpetrators were of opposite sex than the victims of SH.

## Discussion

Female physicians experience the highest rate of SH (74%), with nearly 10% reporting they are verbally sexually harassed more than once per month. However, only 3% of male physicians report being verbally sexually harassed at that frequency. A similar pattern can be observed for physical SH, where female physicians report being physically sexually harassed twice as often as their male counterparts (26% compared to 13%). For female physicians, perpetrators of SH are most often male patients (88% of verbal and 59% physical) followed by male physicians (61% verbal and 48% physical). Male physicians report being sexually harassed most often by female patients (71% verbal and 57% physical).

Among medical students, more than half of women and a third of men were subject to verbal SH. For female medical students, perpetrators of physical SH are most often male physicians followed by male patients. Almost every fourth (24%) female medical student has experienced unwanted physical contact. Prevalence of physical SH is lower for male students (1%), mostly from female patients and male physicians.

The prevalence of sexual harassment according to our survey is higher compared to previous studies conducted in Switzerland. Several factors may explain this discrepancy. On one hand, heightened awareness about sexism and discriminatory behavior may have led to students and physicians reporting SH more often. For example, student collectives against sexual harassment were founded and prevention courses were implemented at multiple Swiss universities, both raising awareness among students and faculty (18–20).

On the other hand, prevalence rates vary depending on the definitions used to describe SH. Neither of the previous studies conducted in Switzerland provided examples of sexually harassing behavior in their questionnaire as we did (6–8), possibly leading to lower prevalence as participants might not identify harassing behavior as sexual harassment.

Lastly, our results are similar to those recently reported by a study conducted in Germany among physicians and nurses in academic medicine, which reports that 74.2% of female physicians have experienced SH during the last 12 months according (21).

Women in our survey consistently reported higher prevalence of SH than men for all frequency categories of both verbal and physical SH. Our analysis showed that the prevalence rates significantly differed as a function of gender. This gender disparity is consistent with previous literature (22,23). Historically male dominated and hierarchical, the health care system upholds a power imbalance, which is thought to facilitate SH (24). However, our survey showed that also a third of men report SH. One possible explanation of comparably high rates among men may be rising awareness and reduced stigma, leading to more men reporting SH. Also, survey anonymity is likely to facilitate the reporting of SH experiences. Future research should include information about the sexuality of male victims and hierarchical rank of perpetrators to identify possible risk factors among men.

Notably, all genders reported patients as the most common perpetrators of SH. Previous studies also found patients and their relatives to be frequent perpetrators of SH, although physicians were cited as the most common perpetrator in most studies (8,21,25). Little is known about why the unique patient-physician relationship seems to be fertile soil for sexually harassing behavior (26). Future research should put additional focus on harassment within the patient-physician relationship and develop strategies to tackle SH specifically by patients.

### Limitations

This study also contains some limitations. There is a possibility of selection bias and over- or underreporting of SH, since we included students from one Swiss university only. However, prevalence among students in our study is similar to a recent study conducted in Germany (27). In addition, SH was not a main purpose of this study and not mentioned in the survey instructions, hence the questions to SH were unlikely to attract a selection of respondents with overrepresented experience of SH.

Secondly, our survey distinguished only between verbal and physical SH, so more subtle forms of harassment in the spectrum of microaggressions might have been missed.

Finally, we failed to recruit participants that identify as gender diverse. However, further research should pay special attention to gender and sexuality minorities as knowing precisely who is at higher risk will be helpful in targeting interventions to tackle SH.

In conclusion, this study provides evidence that the majority of physicians and medical students in Switzerland experience SH by both patients and colleagues. A better understanding of the epidemiology of SH in the current healthcare environment is a much-needed a first step to support the efforts of many physicians and medical students in Switzerland to ultimately establish a respectful and healthy work environment in the medical field (6,7,20,28).

## Acknowledgements

The authors are thankful to the respondents, and to the support by Swiss Society of Internal Medicine, Swiss Young General Practitioners Association (JHaS), Primary Hospital Care and the hospitals involved with distribution of the survey. The authors are thankful to Dr. Kali Tal, University of Bern, for critical review of the manuscript.

## Data availability

The data produced in the present study are available upon reasonable request to the authors.

